# Longitudinal Epidemiology and Variant Dynamics of SARS-CoV-2 in Coastal Kenya (2020–2025): Clinical Features and Wave Patterns

**DOI:** 10.1101/2025.08.22.25334233

**Authors:** Arnold W. Lambisia, Joyce Nyiro, George Githinji, Esther N. Katama, Edidah Moraa, John M. Mwita, Martin Mutunga, Grace Maina, Philip Bejon, My V.T. Phan, Matthew Cotten, Simon Dellicour, L. Isabella Ochola-Oyier, Charles Sande, Edward C. Holmes, James Nyagwange, Charles N. Agoti

## Abstract

**Background:** SARS-CoV-2 is a major cause of outpatient attended acute respiratory infections (ARI). Data from Africa on SARS-CoV-2 infection, variants, symptom profile and longitudinal trends for outpatient presentation is limited.

**Methods:** Starting December 2020, we established an ARI surveillance at five outpatient clinics in coastal Kenya, recruiting ∼15 participants (any age) per week per clinic for SARS-CoV-2 testing and genome analysis. Participants provided respiratory samples, demographic details, vaccination and symptoms data. We compared SARS-CoV-2 clinical and molecular epidemiology pre- and during Omicron waves using multivariable logistic regression.

**Results:** By February 2025, we had recruited 14,562 ARI cases, with 1,053 (7.2%) testing positive for SARS-CoV-2. The median age of cases was 25 years (interquartile range: 15-41) and 65.0% were female. Nine infection waves were recorded with positivity ranging 8.2-25.6%. Inter-wave intervals increased from ≤3 months in 2021 to ≥6 months in 2024. 68 PANGO lineages were identified from 782 (74.2%) sequenced cases, with four predominating local waves (AY.116, BQ.1.8, FY.4.1, LF.7.3.2), rare globally (<0.5%). Overall, common symptoms among positive cases were cough (91.5%), nasal discharge (76.7%) and fever (53.1%). Loss of sense of smell was strongly predictive of COVID-19 in the pre-Omicron era, but body malaise, sore throat, joint pains and nasal discharge were predictive during the Omicron period.

**Conclusion:** SARS-CoV-2 increasingly shows seasonal annual pattern in coastal Kenya, with its clinical features resembling established endemic respiratory viruses. Its case burden is most pronounced in adults. Locally dominant genetic variants may differ from those globally.

**Key points:** Clinical and genomic analysis of SARS-CoV-2 outpatient infections in coastal Kenya (2020–2025) revealed widening inter-wave intervals, repeated predominance of locally distinct PANGO lineages, shifting of predictive symptoms following Omicron emergence underscoring importance of continued local clinical and genomic surveillance.

## Background

By June 15, 2025, over 770 million coronavirus disease 2019 (COVID-19) cases and 7 million associated deaths had been reported to the World Health Organization (WHO) ^1^. This count of COVID-19 cases and deaths is likely a substantial underestimate due to diminished testing and reporting especially after WHO declared the end of the COVID-19 public health emergency of international concern (PHEIC) status in May 2023 ^2^. An understanding of the long-term patterns of SARS-CoV-2 infection waves is only beginning to emerge ^1,3,4^. The virus has sustained recurrent community outbreaks five years into its emergence in human populations, adding a further resource strain on healthcare systems especially in low- and middle-income countries (LMICs) ^5–8^.

Multiple successive waves of SARS-CoV-2 infections, each dominated by a distinct variant have been reported in almost all communities globally ^9^. However, only a limited number of locations have sustained SARS-CoV-2 surveillance beyond the acute pandemic period (2020-2022) to provide insights into emerging variants, wave periodicity, and spatial-temporal lineage dynamics ^10^. In the post-pandemic period, there is a general paucity of data from LMICs on the infection frequency (prevalence), demographics (e.g., age patterns), circulating genetic variants (LMICs contribute ∼0.1% of GISAID data post-pandemic) ^10^, as well as clinical presentation of the infection. This has resulted in a reduced understanding of the ongoing COVID-19 disease burden and clinical presentation ^11^.

Since 2020, COVID-19 clinical presentation has continued to evolve because of changes in circulating variants, existing countermeasures and host immunity ^12–14^. Earlier in the pandemic, loss of the sense of smell and taste were common symptoms among symptomatic COVID-19 cases, helping clinicians identify potential COVID-19 cases and recommend further investigation ^12^. However, with the arrival of the first Omicron variant (lineage BA.1), many groups have described a transition to more typical upper respiratory, systemic, and neurological symptoms in the absence of impaired smell and taste ^13,15,16^. Notably, in sub-Saharan Africa, SARS-CoV-2 infections have largely been asymptomatic (>80%) compared to high income countries (only 22-25% of infections) ^12–17^.

Updating our understanding on SARS-CoV-2 infection and clinical presentation is key in informing clinical diagnosis and planning possible treatment and infection control strategies ^17^. Since December 2020, acute respiratory illness (ARI) surveillance has been ongoing across five selected outpatient health facilities in Kilifi, coastal Kenya ^18^. Leveraging this surveillance platform, we present data on the evolving epidemiological, genomic and clinical profiles of SARS-CoV-2 infections from December 2020 to February 2025 (pre- and during epidemic waves driven by successive Omicron variants).

## Methods

### Study site and selected facilities

The study was conducted within the Kilifi Health and Demographic surveillance system (KHDSS) area, coastal Kenya, leveraging on an ongoing outpatient ARI surveillance platform (**Figure 1**) ^19^. Here, in five outpatient health facilities – Kilifi County Referral Hospital (KCRH, outpatient department), Matsangoni, Mtondia, Mavueni, and Pingilikani – approximately 15 participants presenting with ARI are recruited at each facility per week. The analysis presented here spans the period between December 2020 and February 2025. Participants were recruited if they presented to one of the outpatient facilities with fever (history or measured temperature >38.0°C) and one or more ARI symptoms (see supplementary material) and provided written consent to be involved in the study. New-borns brought to the facilities, individuals with ARI symptoms for more than 28 days, and those who declined consent we excluded from the study.

**Figure 1:**
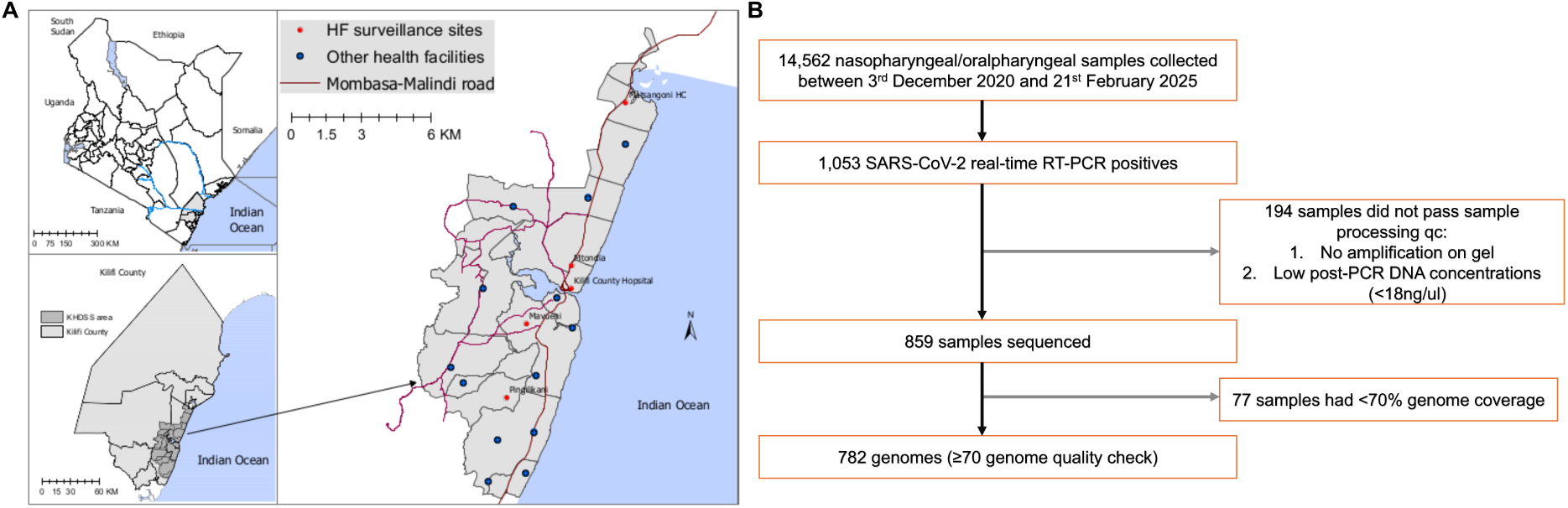
Study map and sample flow chart. **(A)** Map of the study area showing Kilifi Health and Demographic Surveillance System (KHDSS), Kilifi, Kenya and the location of the five health facilities that conducted outpatient surveillance. **(B)** Flow diagram of sample collection and screening from December 2020 and February 2025

### Laboratory procedures

Nasopharyngeal and/or oropharyngeal [NPOP] samples were received at Kenya Medical Research Institute (KEMRI)-Wellcome Trust Research Programme (KWTRP) within 24 hours following collection. Total nucleic acids were extracted and screened for SARS-CoV-2 as previously described ^20^. Briefly, viral ribonucleic acid (RNA) was extracted using either an automated Qiacube HT with a RNeasy extraction kit (Qiagen, Germany) or manually using the QIAamp Viral RNA Mini kit (Qiagen, Germany) and screening done using multiple commercial real-time RT-PCR kits depending on availability during the study period ^21–24^. Samples were defined as positive based on the assay-specific RT-PCR cycle threshold cut-offs and whole genome sequencing was attempted on such samples as described previously on Oxford Nanopore Technologies (ONT) or Illumina MiSeq sequencing platform ^20^. Detailed methods are provided in the supplementary material.

### Genomic data analysis

Raw sequence reads data were assembled to generate consensus genomes using ARTIC bioinformatic pipeline for ONT reads or an inhouse bioinformatic pipeline for Illumina paired-end reads as described previously (https://artic.readthedocs.io/en/latest/) ^5,20^. Using NextClade v.3.10.2 and the data set “nextstrain/sars-cov-2/wuhan-hu-1” v.2025-06-09--15-42-38Z in the command line, PANGO lineages and clades were assigned to the consensus genomes ^25^. Samples whose sequences had <70% genome coverage during sequencing were excluded from downstream analysis due to potential lineage misclassification ^20^.

To assess the relationship between local and global PANGO lineage trends, using GISAID data, we determined if the dominant lineages observed during a local wave in Kilifi were from the top five lineages circulating globally during the corresponding time periods. All genomic data (17.1 million sequences) and associated metadata deposited up to February 28, 2025, were downloaded from GISAID ^10^. The data was subdivided based on time periods of locally observed wave periods and subsampled to achieve 100 sequences per wave, month of collection and country, generating a total of 6,792,560 sequences. The data was then grouped by PANGO lineage to identify the dominant lineages globally across each wave time interval relative to the coastal Kenya patterns.

### Statistical analyses

Demographic data (e.g., date of birth, sex, location/facility), COVID-19 vaccination status, and presenting symptoms were captured using a paper questionnaire by the attending clinicians after interviewing the participant or guardian and transferred to Redcap v.14.3.3 ^26^.

Four outcomes were evaluated: (i) individual SARS-CoV-2 infection status, (ii) SARS-CoV-2 waves of infection, (iii) SARS-CoV-2 variant distribution patterns, and (iv) symptom profiles. The outcomes were analysed over the study period and compared across different periods (pre-Omicron and Omicron epidemic waves). The ‘pre-Omicron period’ spanned between December 2020 and October 2021, while the ‘Omicron period’ began when the first Omicron (BA.1 lineage) cases was observed in the study in November 2021 and extended to February 2025. Weekly trends of SARS-CoV-2 cases and PANGO lineages were summarized across the study period. The positivity rate was estimated within each wave, defined as a period beginning when the weekly positivity rate increased by ≥10% for two consecutive weeks and ending when the positivity rate declined for two weeks and returned to within 5% of the wave starting level ^27–29^.

To compare age, sex and vaccination status between SARS-CoV-2 positive and negative individuals a chi-square test was used. Multivariable logistic regression models were constructed to assess associations between individual symptoms and SARS-CoV-2 infections adjusting for age, sex, and study period. The model equation was as follows:

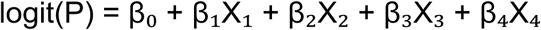

where:

β₀ = intercept

β₁, β₂, β₃, β₄ = regression coefficients

P represents the probability of SARS-CoV-2 infection status (0 = negative, 1 = positive)

X₁ = individual symptom (No, Yes)

X₂ = age (continuous variable, years)

X₃ = sex (female, male)

X₄ = study period (Pre-Omicron and Omicron)

Separate models were built for the pre-Omicron and Omicron periods, in addition to an overall model spanning the entire study. Age, sex and period (pre-Omicron and Omicron) were adjusted for in the models. Results are reported as adjusted odds ratios (aOR), and 95% confidence intervals (CI) were reported with p-values less than 0.05 considered statistically significant. Cases with missing symptom data were removed before performing multivariable analysis. All data processing and statistical analyses were conducted in Python v3.13 and R v4.2.

## Results

Between December 3, 2020, and February 21, 2025, a total of 14,562 NPOP samples were collected from patients presenting with ARI at five outpatient health facilities in coastal Kenya (**Table 1**). Of these, 1053 (7.2%) samples were positive for SARS-CoV-2. There was a significant difference in the distribution of SARS-CoV-2 cases across different study years (*p* < 0.001) and facilities (*p* = 0.002; **Table 1**). Among the 1,053 positives, 859 (81.5%) passed sample processing quality control and were sequenced. 77 sequenced samples with genome coverage <70% coverage were dropped from further lineage analyses (**Figure 1**). The median Ct value was significantly lower for the sequenced (23.1, interquartile range [IQR] = [19.6-26.7]) compared to those with low genome coverage (28.4, IQR = [26.4-29.9]) and those samples not sequence (33.2, IQR = [31.6-34.9]; p <0.001).

**Table 1:** Baseline characteristics of SARS-CoV-2 positive and negative cases presenting with ARI in five outpatient facilities in the KHDSS between December 2020 and February 2025.

|  | Positive<br>(n=1053) | Negative<br>(n=13509) | Total (n=14562) | *p value* |
| --- | --- | --- | --- | --- |
| **Sex** |  |  |  | 0.037 |
| Female | 684 (65.0%) | 8336 (61.7%) | 9020 (61.9%) |  |
| **Age years** |  |  |  |  |
| Median (IQR) | 25 (15 - 41) | 17 (7 - 33) | 18 (7 - 34) |  |
| **Age group** |  |  |  | < 0.001 |
| 0-4 | 106 (10.1%) | 2553 (18.9%) | 2659 (18.3%) |  |
| 5-9 | 66 (6.3%) | 1643 (12.2%) | 1709 (11.7%) |  |
| 10-19 | 210 (19.9%) | 3338 (24.7%) | 3548 (24.4%) |  |
| 20-39 | 372 (35.3%) | 3170 (23.5%) | 3542 (24.3%) |  |
| 40-64 | 230 (21.8%) | 2038 (15.1%) | 2268 (15.6%) |  |
| 65+ | 69 (6.6%) | 767 (5.7%) | 836 (5.7%) |  |
| **Year of collection** |  |  |  | <0.001 |
| 2020 | - | 4 (0.0%) | 4 (0.0%) |  |
| 2021 | 351 (33.3%) | 2501 (18.5%) | 2852 (19.6%) |  |
| 2022 | 353 (33.5%) | 3275 (24.2%) | 3628 (24.9%) |  |
| 2023 | 181 (17.2%) | 3566 (26.4%) | 3747 (25.7%) |  |
| 2024 | 159 (15.1%) | 3624 (26.8%) | 3783 (26.0%) |  |
| 2025 | 9 (0.9%) | 539 (4.0%) | 548 (3.8%) |  |
| **Health facility** |  |  |  | 0.002 |
| Kilifi County Hospital | 247 (23.5%) | 3074 (22.7%) | 3321 (22.8%) |  |
| Matsangoni | 195 (18.5%) | 2569 (19.0%) | 2764 (19.0%) |  |
| Mavueni | 179 (17.0%) | 2686 (19.9%) | 2865 (19.7%) |  |
| Mtondia | 241 (22.9%) | 2472 (18.3%) | 2713 (18.6%) |  |
| Pingilikani | 191 (18.1%) | 2708 (20.0%) | 2899 (19.9%) |  |
| **Vaccination status<sup>#</sup>** |  |  |  |  |
| Yes | 141 (13.4%) | 1552 (11.5%) | 1693 (11.6%) |  |
| No | 587 (55.7%) | 9803 (72.6%) | 10390 (71.4%) |  |
| No data | 325 (30.9%) | 2154 (15.9%) | 2479 (17.0%) |  |

SARS-CoV-2 positive and negative cases differed in their sex distribution (female; 65.0% and 61.7%, respectively; *p* = 0.037) and age (median 25 vs 17 years; *p* <0.001) as shown in **Table 1**. The age groups 20-39 (35.3% vs 23.5%), 40-64 (21.8% vs 15.1%) and ≥65 (6.6% vs 5.7%) years also had a greater proportion of positive than negative cases (**Table 1**).

The first SARS-CoV-2 case in Kenya was reported on March 12, 2020. Since that time ten waves of infection have been observed in Kilifi, Kenya, although the first epidemic wave recorded few infections and exhibited limited spread ^22,23,30–32^. Here, we analysed data collected between December 3, 2020, to February 21, 2025,2025, spanning nine waves of infection (**Figure 2A)**. During wave periods, SARS-CoV-2 positivity rate ranged from 8.2% (95% CI = [6.5-10.1%]) to 25.6% (95% CI = [22.6-28.7%]). In contrast, in non-wave periods the prevalence was 0.6% (95% CI = [0.4-0.8%]; **Figure 2C**). In 2021, four waves of infection were observed. These were dominated by the variants B.1.530, B.1.1.7 (Alpha), AY.116 (Delta) and BA.1.1 (Omicron), respectively, with the time intervals between them ranging from two weeks to three months. In 2022 and 2023, the time interval between consecutive waves gradually increased to approximately five months and by 2024-2025, waves were observed nine months apart. The average duration of waves was 2.8 months (range: 1.2 to 3.7 months).

**Figure 2:**
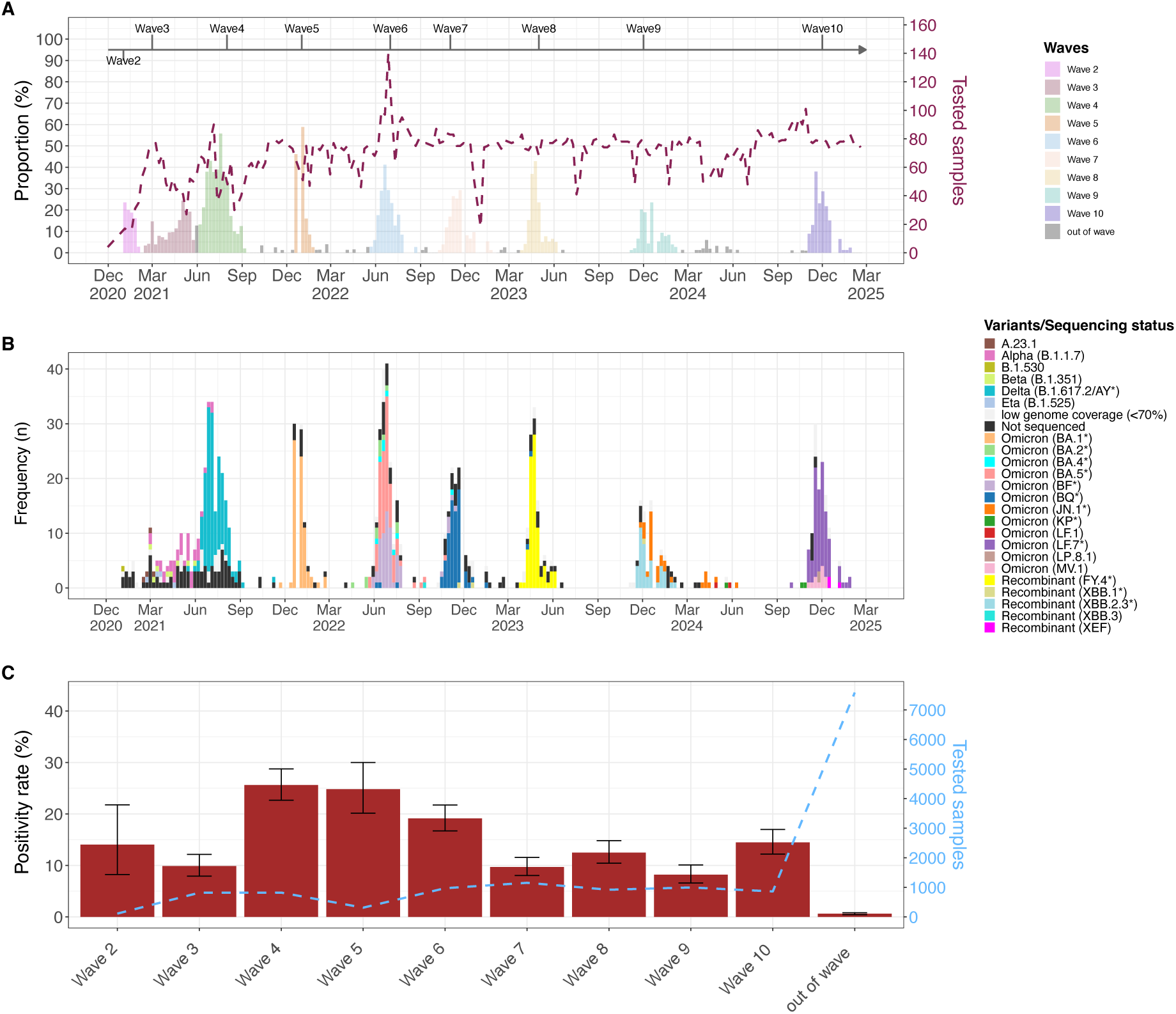
Temporal trends SARS-CoV-2 cases, wave positivity rates and PANGO lineage. **(A)** Weekly temporal trends of SARS-CoV-2 cases from December 2020 and February 2025 in five outpatient facilities with the KHDSS in Kilifi, Kenya. The dashed brown line shows the number of samples tested per week (secondary y-axis) and the bars show the weekly proportion of tested samples that were positive are coloured by waves. **(B)** Weekly temporal trends of SARS-CoV-2 variants identified during the study period. **(C)** Positivity rate of SARS-CoV-2 across different waves and out of wave. The dashed blue line shows the number of samples tested per wave (secondary y-axis).

### Distribution of lineages across waves

Between January 8, 2021, and January 28, 2025, 68 PANGO lineages were identified from the 782 sequenced cases (**Figure 2B**; **Supplementary figure 1**). A median of 7 (IQR = [5-9]) PANGO lineages were detected in each wave with one to two predominant lineages (**Figure 3**). In waves 3 and 5, similar lineages were dominant locally and globally; i.e., Alpha (B.1.1.7; local = 64%, global = 51%) and Omicron (BA.1.1, 82% vs 30.4%). During wave 6, there was joint dominance of two lineages locally; BA.5.2.1 (35%) and BF.9 (an alias of BA.5.2.1.9; 32.4%), while at a global level BA.5.2.1 (7.3%) was among the top circulating lineages while BF.9 was less common (0.01%). Similarly, during wave 9, there was joint dominance of XBB.2.3-derived lineages (KT.1 [31.7%], KH.1 [12.6%] and GE.1.2 [9.5%]) and JN.1-like (JN.1; 17.4% and JN.1.4.7; 11.1%) lineages, comprising 82.3% of the sequenced genomes, whereas globally only JN.1 was the top lineage detected (21.6%). During this wave, the XBB.2.3-like lineages comprised less than 0.01% of the global lineages. During waves 4, 7, 8 and 10, B.1.617.2 (14.1%), BQ.1.1 (alias BA.5.3.1.1.1.1.1.1, 13.5%), XBB.1.5 (16.9%) and XEC (37.5%) were the top lineages detected globally, while AY.116 (77.1%), BQ.1.8 (76.2%), FY.4.1 (alias XBB.1.22.1.4.1, 85.1%) and LF.7.3-like (61.4%) were dominant locally (**Figure 3, Supplementary figure 2**). Notably, the dominant local lineages during these four waves, AY.116, BQ.1.8, FY.4.1 and LF.7.3-like, comprised <0.5% of the globally reported sequences during the same period.

**Figure 3:**
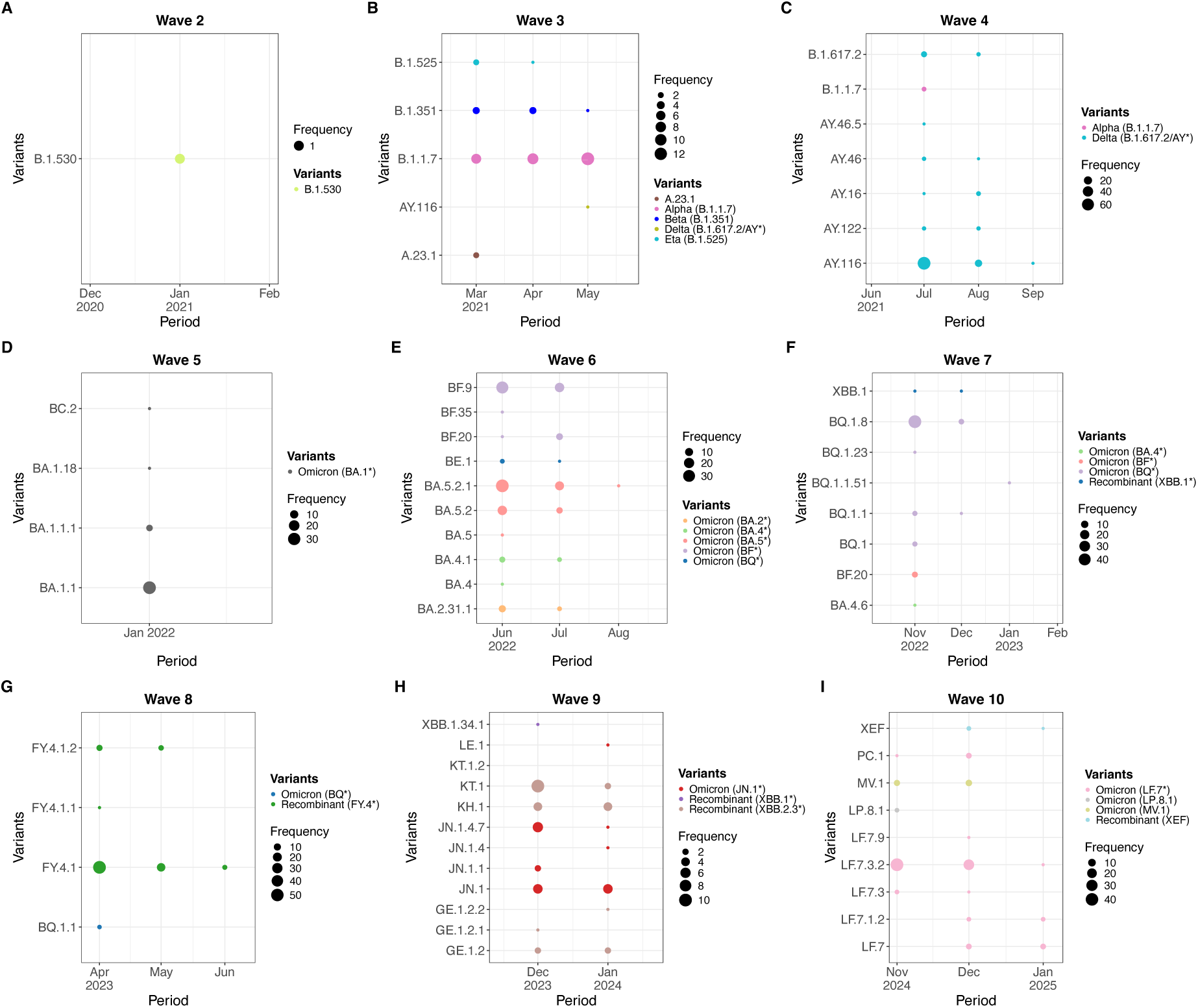
Temporal patterns of PANGO lineages across different waves in Kilifi Kenya between December 2021 and February 2025. The PANGO lineages are grouped by wave and plotted on the y-axis with time on the x-axis. The lineages are coloured by variants in the legend.

### Clinical presentation of SARS-CoV-2 positive cases

During the study period, the most prevalent symptom among SARS-CoV-2 positive individuals seeking outpatient care was cough (*n* = 964; 91.5%), followed by nasal discharge (*n* = 808; 76·7%), fever (*n* = 559; 53·1%), joint pains (*n* = 337; 32·0%), body malaise (*n* = 332; 31·5%), and sore throat (*n* = 312; 29·6%; **Supplementary Figure 3**). The most reported symptoms were consistent across both the pre-Omicron and Omicron periods (**Supplementary Figure 3A**). Individuals reported a median of three symptoms (range: 1 to 10) during a clinic visit (**Supplementary Figure 3B**). The proportion of individuals presenting by each symptom by age group was calculated across age groups ≥5 years. We did not include the <5-year-olds as some symptoms (e.g., headache) are difficult to capture for this age group. Cough, chest pain, crackles and difficulty in breathing were more prevalent in individuals 65 years and above (**Supplementary Figure 3C)**.

Using a multivariable logistic regression model adjusted for age, sex and period, we investigated the association between SARS-CoV-2 infection status and individual symptoms over the study period. From this, we observed that headache (aOR = 1.58; 95% CI = [1.33-1.87]; *p* < 0.001), body malaise (aOR = 1.35; 95% CI = [1.17-1.55]; p < 0.001), loss of smell (aOR = 6.11; 95% CI = [2.41-14.96]; *p* < 0.001), nasal discharge (aOR = 1.33; 95% CI = [1.15-1.55]; *p* < 0.001), and joint pains (aOR = 1.16; 95% CI = [1.01-1.33]; p = 0.03) were strongly associated with SARS-CoV-2 infection (**Figure 4A**). In contrast, fever, crackles, wheezes, epigastric pain and difficulty in breathing were negatively associated with a SARS-CoV-2 infection (**Figure 4A**).

**Figure 4:**
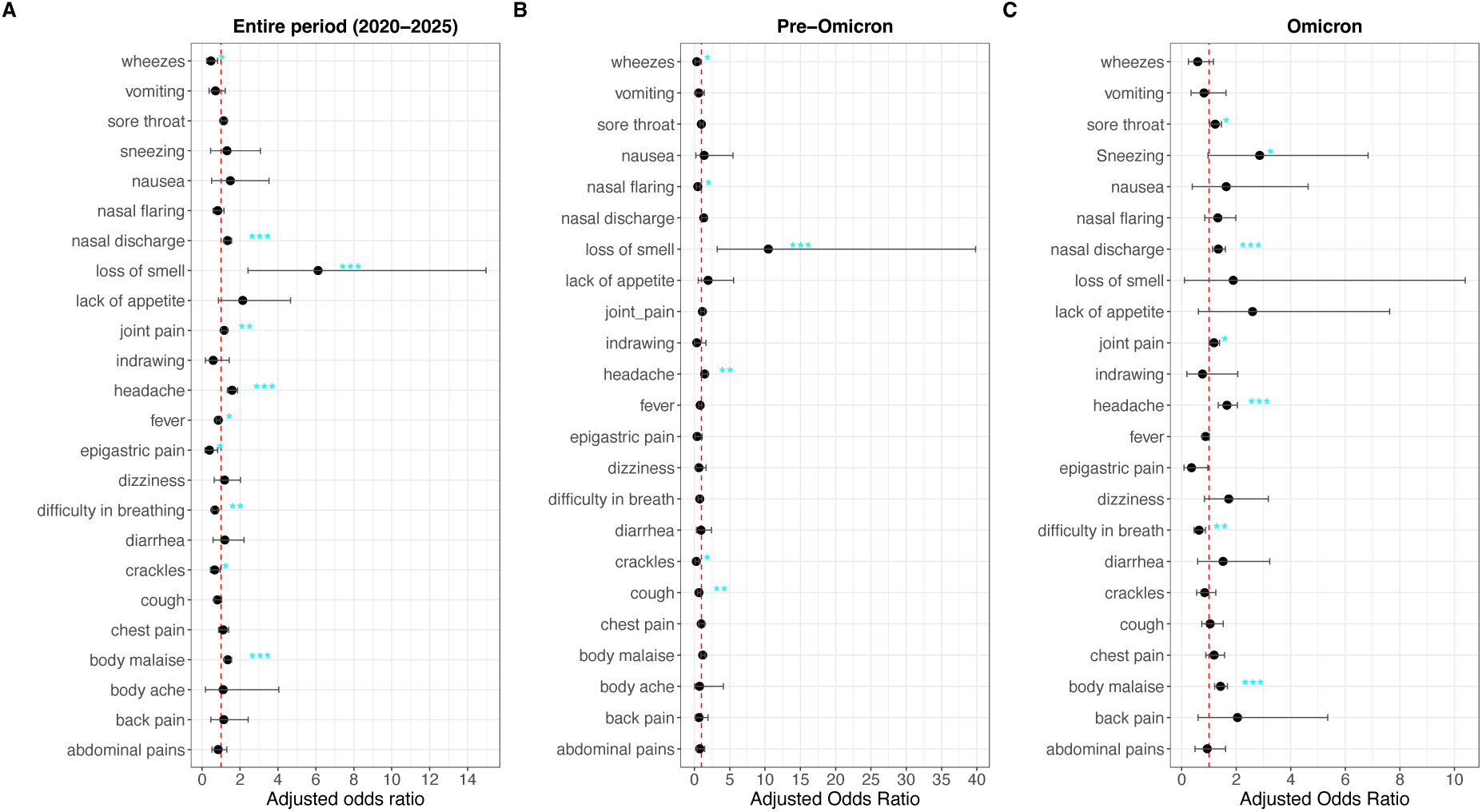
Adjusted odds ratio showing association between SARS-CoV-2 infection status and individual symptoms across **(A)** the entire study period, **(B)** pre-Omicron (December 2020 to October 2021) and **(C)** during Omicron (November 2021 to February 2025) periods.

When stratified by period, headache (aOR = 1.45; 95% CI = [1.08-1.92]; *p* = 0.009) and loss of smell (aOR = 10.46; 95% CI = [3.22-39.82]; p < 0.001) were significantly associated with a SARS-CoV-2 infection during the pre-omicron period (**Figure 4B**). During the Omicron period, headache (aOR = 1.65; 95% CI = [1.33 to 2.04]; *p* < 0.001), body malaise (aOR = 1.42; 95% CI = [1.20-1.67]; *p* < 0.001), nasal discharge (aOR = 1.34; 95% CI = [1.13-1.60]; *p* < 0.001), sore throat (aOR = 1.23; 95% CI = [1.03-1.46]; *p* = 0.017), and joint pains (aOR = 1.18; 95% CI = [1.00 to 1.39]; *p* = 0.045) were highly associated with SARS-CoV-2 infection (**Figure 4C**).

The Omicron lineages categorized into four groups (**Supplementary Table 1**) – Omicron-BA.1/2, Omicron-BA.4/5, Omicron-XBB (a recombinant variant) and Omicron-BA.2.86.1 (a.k.a. JN.1) – showed a significant difference in distribution by age group. Compared to other Omicron variants, XBB was more prevalent among children below 5 years of age and adults above 65 years. The occurrence of the symptom of body malaise, sore throat, headache, chest pain, fever, cough, nasal discharge, nasal flaring, dizziness and joint pains differed significantly across different Omicron variants (**Supplementary Table 2**).

Compared to Omicron-BA.2.86.1, Omicron-BA.1/2 was strongly associated with joint pains (aOR = 2.83; 95% CI = [1.58-5.12]; *p* < 0.001), headache (aOR = 2.84; 95% CI = [1.44-5.71]; *p* = 0.002), body malaise (aOR = 2.07; 95% CI = [1.16-3.71]; *p* = 0.01), sore throat (aOR = 3.94; 95% CI = [2.11-7.48]; *p* < 0.001), nasal discharge (aOR = 2.29; 95% CI = [1.20-4.56]; *p* = 0.014), and nasal flaring (aOR = 5.26; 95% CI = [1.17-36.62]; *p* < 0.001). Omicron BA.4/5 was strongly associated with body malaise (aOR = 1.60; 95% CI = [1.01-2.57]; *p* < 0.001), sore throat (aOR = 1.99; 95% CI = [1.17-3.47]; *p* = 0.01), chest pain (aOR = 8.03; 95% CI = [2.79-33.97]; *p* < 0.001) and nasal discharge (aOR = 1.86; 95% CI = [1.15-3.02]; *p* = 0.01). XBB-like variants were strongly associated with sore throat (aOR = 2.05; 95% CI = [1.14-3.75]; *p* = 0.017) and cough (aOR = 8.95; 95% CI = [1.64-166.39]; *p* = 0.039). No other symptoms differed significantly across variants.

## Discussion

We describe the molecular and clinical epidemiology of COVID-19 over multiple waves of infection in coastal Kenya between December 2020 and February 2025. Such a longitudinal view of COVID-19 prevalence, lineage dynamics and symptom profiles during the pre-Omicron and Omicron periods are rare in African settings ^33^. Across nine waves of infections, we determined that within-wave test-positivity rate ranged from 8 to 26%. Overall, 7.2% of tested participants were SARS-CoV-2 positive. Detailed genomic analysis within these waves has been previously presented for all but wave 10 ^23,34–36^. Here we include wave 10 and have examined the longitudinal trends of all our data to date.

Our data revealed that most SARS-CoV-2 infections occurred in adolescents and young adults, a pattern distinct from other viral respiratory infections which typically affect children under five predominantly ^37^. The highest positivity rate within waves was observed during the Delta (wave 4; 25.6%) and Omicron-BA.1 (wave 5; 24.8%) waves. This likely reflect a combination of factors: the immune vulnerability of the population, and the variants’ ability to spread, cause disease, and evade immune responses ^33,38^. Following this, there was a gradual decline in positivity rate, potentially reflecting the growing and high pre-existing population immunity in the local population with each new wave^39,40^. The Delta variant of concern (VOC) has previously been reported to have increased odds of causing severe disease and hospitalizations compared to the ancestral, Alpha and even Omicron variants ^41,42^, potentially explaining the high positivity rate during wave 4 dominated by the Delta VOC in Kilifi, Kenya, when the seroprevalence rate was 24.7% (95% CI: 17.5– 32.6%) at the start of the wave ^43^.

Notably, the time interval between waves has been increasing over time from a few weeks to near annually in 2024-2025, suggesting that the SARS-CoV-2 epidemics could be settling to a seasonal annual epidemic pattern, although longer surveillance is required to confirm this. In the local population, reactive IgG antibodies that could neutralize the ancestral virus *in vitro* were reported in 42% of pre-pandemic samples ^44^ . However, over time, the SARS-CoV-2 seroprevalence grew to 77.4% by May 2022 in coastal Kenya, with limited neutralizing immune responses reported among emerging Omicron subvariants ^39,40^. Therefore, it appears for a new wave to be observed, both significant previous responses waning and introduction a new variant with sufficient immune escape, high infectivity and transmissibility area required.

Our previous work found that the observed lineages dominating different waves were introduced through international and local human mobility routes in the region, with disparities in the locally and global dominating variants in four waves (i.e. waves 4, 7, 8 and 10). This indicates that SARS-CoV-2 variant dominance global may not necessarily predict local epidemics as there is region specific SARS-CoV-2 variant dominance and replacement. In turn, this emphasizes the need for sustained local genomic surveillance to inform data driven public health responses. Further, variants including FY.4.1, predominant in wave 7, have been shown to have emerged locally and then spread globally, such that some global waves can be driven by locally emerging variants ^45^.

Like studies conducted elsewhere ^12,15–17^, we observed that some clinical symptoms are predictive of COVID-19 disease and there has been a shift in SARS-CoV-2 clinical presentation during the pre-Omicron to Omicron periods. Early in the pandemic, infections with ancestral lineages presented frequently, with symptoms including loss of the senses of smell and taste, and fatigue in ARI cases. This was unlike Delta VOC infections which largely presented with moderate to severe lower respiratory tract symptoms, causing millions of fatalities, while Omicron infections present with mild upper respiratory symptoms and systemic symptoms ^12,13,41,46^. This has made SARS-CoV-2 infections increasingly indistinguishable from other common respiratory viruses ^47^ (ref).

Among Omicron subvariants, including BA.1, BA.2, BA.5, and XBB, differences in clinical profiles have been described as observed here ^15,16^. The emergence of Omicron lineages with a BA.2.86.1 backbone (a.k.a. JN.1) led to an upsurge of cases globally and, in this study, we observed that compared to these variants; Omicron BA.1/2 were strongly associated with joint pains, headache, body malaise, sore throat, nasal discharge and nasal flaring, Omicron-BA.4/5 were strongly associated with body malaise, sore throat, chest pain and nasal discharge and XBB-like variants were strongly associated with sore throat and cough. This finding shows that routine linkage of syndromic surveillance with genomic data is critical in understanding the evolving infection clinical presentation, informing formulation of guidelines on case definitions, clinical diagnosis and management by clinicians.

This study had limitations. First, we only captured only a fraction of symptomatic ARI cases in five outpatient clinics (∼15 cases per week) and did not include mildly symptomatic or asymptomatic cases which make the greatest fraction of COVID-19 cases in Africa ^11^. Our previous work based on community surveillance indicated that up to 83% of the SARS-CoV-2 infections were asymptomatic ^21^. Second, facility-based surveillance is biased by the background population health seeking behaviour which may have varied since the pandemic started, and there may also be gender biases. Third, we have not assessed the SARS-CoV-2 immune status of the individuals we analysed, which may influence the COVID-19 clinical profile. Nonetheless, our findings are largely consistent with previous studies globally in highlighting the evolution of the clinical profile of SARS-CoV-2 infections.

In conclusion, using an out-patient surveillance platform, we show how SARS-CoV-2 wave dynamics have substantially evolved on the Kenyan coast over the past five years. Distinct PANGO lineages dominated different waves, some clearly introduced from other global locations others unique and potentially first occurring locally. The clinical profiles have changed from the pre-Omicron to Omicron periods and can be distinct even among different Omicron subvariants. Sustained sentinel surveillance that links syndromic and genomic surveillance is crucial is critical in identifying emerging SARS-CoV-2 variants, updating knowledge on SARS-CoV-2 clinical profile, and predicting the nature and timing of future waves in the population.

## Acknowledgement

We are grateful to the study participants who provided samples and members of the Pathogen Epidemiology and Omics (PEO) group at KEMRI-Wellcome Trust Research Programme who did sample collection (field team) and laboratory processing (laboratory team). We thank Prof. D. James Nokes (retired) who lead the study investigations 2020-2023. We are grateful to Mr. Christopher Nyundo who helped with generating the study area map. This work is published with permission from director KEMRI.

## Members of the Pathogen Epidemiology and Omics Team

### Field Team

Swabah Salim, Erick Shoboi, Eunice Nyiro, Leah Mpathe, Racheal Nyamvula, Timothy Mwaringa, Celian Kamandi, Phanice Rongoma, Ismile Mwamuye, Elena Dena, Justin Kadhengi, Oscar Mwasambu, Loice Masha, Donald Deche, Lucy Charo, Phanice Amukhale, Emily Nyale and Joan Omungala

### Lab team

Robinson Cheruiyot, Clement Lewa, Simon Mutula and Hillary Wafula

## Ethical approval

The protocols consenting and sample collection process were reviewed and approved by KEMRI Scientific Ethics and Research Unit (SERU), Nairobi Kenya (protocol number #3103). Samples (nasopharyngeal and/or oropharyngeal swabs) were collected following consent from a parent or guardian for participants aged <18-year-olds (with assent for children aged between 13–18-year-old). Individual written informed consent was sought for participants aged >18 years.

## Author contributions

AWL, EM, JMM and MM were involved in investigation. CNA, GG and JUN acquired the funds for the study and conceptualized the study. E.K was involved in data curation. CNA, GG, JN and CS were involved in supervision. AWL and CNA wrote the original draft, reviewed and edited the manuscript. PB, GG, SD, MVTP, MC, LIO, ECH were involved in writing – reviewing and editing. AWL and EK were involved in data curation and visualization. All authors reviewed the manuscript.

## Data availability

The final consensus genomes from the SARS-CoV-2 samples sequenced in this study have been deposited in the Global Initiative on Sharing all Influenza Data (GISAID) database and can be accessed at https://doi.org/10.55876/gis8.250723bc48. Epidemiological data and scripts for data analysis are available on the Harvard dataverse ^49^.

