## supplementary tables and figures for "Longitudinal Epidemiology and Variant Dynamics of SARS-CoV-2 in Coastal Kenya (2020–2025): Clinical Features and Wave Patterns"

[Supplementary Figure 3: Symptom epidemiology in SARS-CoV-2 positive individuals seeking outpatient care. **(A)** Prevalence of COVID-19 symptoms in the pre-Omicron and Omicron periods. Error bars indicate 95% credible intervals (Cis). **(B**) Frequency of symptoms reported per individual. The dotted line shows the median number of symptoms reported per individual. **(C)** Prevalence of COVID-19 symptoms by age. A cut-off of five years and above was used to avoid bias among the symptoms as some cannot be recorded for children (e.g., headache). A gradient colour scale is used to show high and low proportions.	9](#_Toc206767075)

### Supplementary Material and Methods

#### SARS-CoV-2 screening

To screen for SARS-CoV-2, the following kits/protocols were used: (1) Sansure Biotech Novel Coronavirus (2019-nCoV) Nucleic Acid Diagnostic real-time RT-PCR kit, (2) TaqPath™ COVID-19 Fast PCR Kit (N gene), (3) European Virus Archive – GLOBAL (EVA-g) primers (targeting E or RdRp or ORF1AB genes) and (4) an in-house real-time RT-PCR with primers/probe targeting the envelope (E) gene (forward: 5'- ACA GGT ACG TTA ATA GTT AAT AGC GT -3', reverse: 5'- ATA TTG CAG CAG TAC GCA CAC A -3' and probe 5'-ACA CTA GCC ATC CTT ACT GCG CTT CG-3') and QIAGEN Multiplex RT-PCR + R Kit (QIAGEN, UK). Positives were determined based on kit/protocol determined cut-offs ^1,2^. For quality control, negative and positive controls were included both at the extraction and RT-PCR stages.

#### SARS-CoV-2 whole genome sequencing (WGS)

Ribonucleic acids (RNA) were re-extracted from 140µl of positive samples using the QIAamp Viral RNA Mini Kit (QIAGEN, UK). 8ul of RNA was reverse-transcribed using 2ml of LunaScript RT Mix (NEB, E3010, MA, USA) by incubating at 25◦C for 2 min, 55 °C for 10 min and 95 °C for 10 min. The cDNA was the PCR amplified in 2 pools by combining 1.3ul of cDNA, 2μl of primer pool, 6.3μl of Q5 Hot Start High-Fidelity 2X Master Mix (NEB M0494, MA, USA) and 1.9μl of nuclease-free water ^2,3^. The thermocycling conditions were: 1 cycle of 98°C for 30 s, followed by 25 cycles of 98 °C for 30 s and 65 °C for 5 minutes, 15 cycles of 62.5 °C for 5 minutes and 98 °C for 15 seconds and 1 cycle of 62.5 °C for 5 minutes. The two reactions were pooled, cleaned using 1X AMPure XP beads (Beckman Coulter, A63881, Indianapolis, USA). An *ad hoc* quality control strategy described previously was used to exclude samples with low concentrations (<28 ng/ml) ^3^. Libraries were prepared as per the ARTIC SARS-CoV-2 sequencing protocol ^4^ and sequenced on the Oxford Nanopore GridION or using the COVIDSeq Assay (Illumina US) and sequenced on the Illumina MiSeq platform.

#### Presenting symptoms

Presence or absence of body malaise, sore throat, headache, chest pain, fever, cough, nasal discharge, loss of smell, lack of appetite, difficulty in breathing, nasal flaring, crackles, abdominal pains, dizziness, vomiting, wheezes, diarrhoea, back pain, joint pain, sneezing, epigastric pain, nausea, indrawing, body ache and feed unable.

#### Supplementary Table 1: Grouping of SARS-CoV-2 lineages into variants

| Variant |  |  | PANGO Lineages |
| --- | --- | --- | --- |
| Ancestral | – | _ | B.1.530 & A.23.1 |
| Alpha | – | Alpha (B.1.1.7) | B.1.1.7 |
| Beta | – | Beta (B.1.351) | B.1.351 |
| Delta | – | Delta (B.1.617.2/AY*) | AY.116, AY.122, AY.16, AY.46, AY.46.5 & B.1.617.2 |
| Omicron | Omicron BA.1/2 | Omicron (BA.1*) | BA.1, BA.1.1, BA.1.1.1, BA.1.1.4, BA.1.18 & BC.2 |
|  |  | Omicron (BA.2*) | BA.2 & BA.2.31.1, |
|  | Omicron BA.4/5 | Omicron (BA.4*) | BA.4, BA.4.1 & BA.4.6, |
|  |  | Omicron (BA.5*) | BA.5, BA.5.2 & BA.5.2.1 |
|  |  | Omicron (BQ*) | BQ.1, BQ.1.1, BQ.1.1.51, BQ.1.23, BQ.1.8 & BE.1 |
|  |  | Omicron (BF*) | BF.20, BF.35 & BF.9, |
|  | Omicron BA.2.86.1 | Omicron (JN.1*) | JN.1, JN.1.1, JN.1.16.1, JN.1.18, JN.1.4, JN.1.4.7, LE.1, LE.1.1 & LE.1.3 |
|  |  | Omicron (LF.1) | LF.1 |
|  |  | Omicron (LF.7*) | LF.7, LF.7.1.2, LF.7.3, LF.7.3.2 & LF.7.9, |
|  |  | Omicron (KP*) | KP.2.3, KP.3.1.1 LP.8.1, MV.1 & PC.1 |
|  |  | Omicron (MV.1*) | MV.1 |
|  |  | Omicron (LP.8) | LP.8.1 |
| Recombinant | Recombinant | Recombinant (FY.4*) | FY.4.1, FY.4.1.1 & FY.4.1.2 |
|  |  | Recombinant (XBB.2.3*) | GE.1.2, GE.1.2.1, GE.1.2.2, GS.4.1, KH.1, KT.1 & KT.1.2, |
|  |  | Recombinant (XEF) | XEF |
|  |  | Recombinant (XBB.1*) | XBB.1 & XBB.1.3.1, |
|  |  | Recombinant (XBB.3 | XBB.3 |
| VOI | – | Eta (B.1.525) | B.1.525 |

#### Supplementary Table 2: Comparison of demographic characteristics and individual symptoms across different Omicron variants from health facility surveillance in Kilifi, Kenya.

|  | Omicron BA.1/2 (n=84) | Omicron BA.4/5 (n=224) | XBB (n=132) | Omicron BA.2.86.1 (n=138) | Total (n=578) | *p value* |
| --- | --- | --- | --- | --- | --- | --- |
| Sex |  |  |  |  |  | **0.185** |
| Female | 52 (61.9%) | 166 (74.1%) | 89 (67.4%) | 95 (68.8%) | 402 (69.6%) |  |
| Age Group |  |  |  |  |  | **< 0.001** |
| 0 - 4 | 5 (6.0%) | 13 (5.8%) | 29 (22.0%) | 12 (8.7%) | 59 (10.2%) |  |
| 5 - 9 | 11 (13.1%) | 11 (4.9%) | 9 (6.8%) | 12 (8.7%) | 43 (7.4%) |  |
| 10 - 19 | 9 (10.7%) | 62 (27.7%) | 21 (15.9%) | 23 (16.7%) | 115 (19.9%) |  |
| 20 - 39 | 35 (41.7%) | 82 (36.6%) | 37 (28.0%) | 51 (37.0%) | 205 (35.5%) |  |
| 40 - 64 | 23 (27.4%) | 48 (21.4%) | 24 (18.2%) | 31 (22.5%) | 126 (21.8%) |  |
| 65+ | 1 (1.2%) | 8 (3.6%) | 12 (9.1%) | 9 (6.5%) | 30 (5.2%) |  |
| Clinical presentation |  |  |  |  |  |  |
| Body malaise | 36 (42.9%) | 82 (36.6%) | 36 (27.3%) | 37 (26.8%) | 199 (33.0%) | **0.025** |
| Sore throat | 36 (42.9%) | 63 (28.1%) | 37 (28.0%) | 23 (16.7%) | 159 (27.5%) | **< 0.001** |
| Headache | 25 (29.8%) | 47 (21.0%) | 14 (10.6%) | 18 (13.0%) | 104 (18.0%) | **< 0.001** |
| Chest pain | 6 (7.1%) | 33 (14.7%) | 1 (0.8%) | 3 (2.2%) | 43 (7.4%) | **< 0.001** |
| Fever | 43 (51.2%) | 112 (50.0%) | 96 (72.7%) | 88 (63.8%) | 339 (58.7%) | **< 0.001** |
| Cough | 82 (97.6%) | 210 (93.8%) | 131 (99.2%) | 129 (93.5%) | 552 (95.5%) | 0.044 |
| Nasal discharge | 69 (82.1%) | 177 (79.0%) | 98 (74.2%) | 92 (66.7%) | 436 (75.4%) | **0.024** |
| Lack of appetite | 0 (0.0%) | 1 (0.4%) | 0 (0.0%) | 0 (0.0%) | 1 (0.2%) | 0.663 |
| Difficulty breathing | 4 (4.8%) | 11 (4.9%) | 7 (5.3%) | 4 (2.9%) | 26 (4.5%) | 0.771 |
| Nasal flaring | 6 (7.1%) | 3 (1.3%) | 4 (3.0%) | 2 (1.4%) | 15 (2.6%) | **0.028** |
| Crackles | 4 (4.8%) | 11 (4.9%) | 4 (3.0%) | 0 (0.0%) | 19 (3.3%) | 0.067 |
| Abdominal pains | 2 (2.4%) | 2 (0.9%) | 0 (0.0%) | 3 (2.2%) | 7 (1.2%) | 0.279 |
| Dizziness | 1 (1.2%) | 7 (3.1%) | 0 (0.0%) | 0 (0.0%) | 8 (1.4%) | **0.032** |
| Vomiting | 1 (1.2%) | 2 (0.9%) | 0 (0.0%) | 1 (0.7%) | 4 (0.7%) | 0.716 |
| Wheezes | 0 (0.0%) | 3 (1.3%) | 1 (0.8%) | 0 (0.0%) | 4 (0.7%) | 0.404 |
| Diarrhea | 1 (1.2%) | 0 (0.0%) | 0 (0.0%) | 1 (0.7%) | 2 (0.3%) | 0.315 |
| Backpain | 0 (0.0%) | 3 (1.3%) | 0 (0.0%) | 0 (0.0%) | 3 (0.5%) | 0.19 |
| Joint pains | 39 (46.4%) | 71 (31.7%) | 36 (27.3%) | 33 (23.9%) | 179 (31.0%) | 0.004 |
| Sneezing | 1 (1.2%) | 1 (0.4%) | 2 (1.5%) | 0 (0.0%) | 4 (0.7%) | 0.43 |
| Epigastric pain | 2 (2.4%) | 1 (0.4%) | 0 (0.0%) | 0 (0.0%) | 3 (0.5%) | 0.07 |
| Nausea | 0 (0.0%) | 2 (0.9%) | 0 (0.0%) | 0 (0.0%) | 2 (0.3%) | 0.366 |
| Indrawing | 0 (0.0%) | 2 (0.9%) | 0 (0.0%) | 0 (0.0%) | 2 (0.3%) | 0.366 |
| Body ache | 0 (0.0%) | 1 (0.4%) | 0 (0.0%) | 0 (0.0%) | 1 (0.2%) | 0.663 |

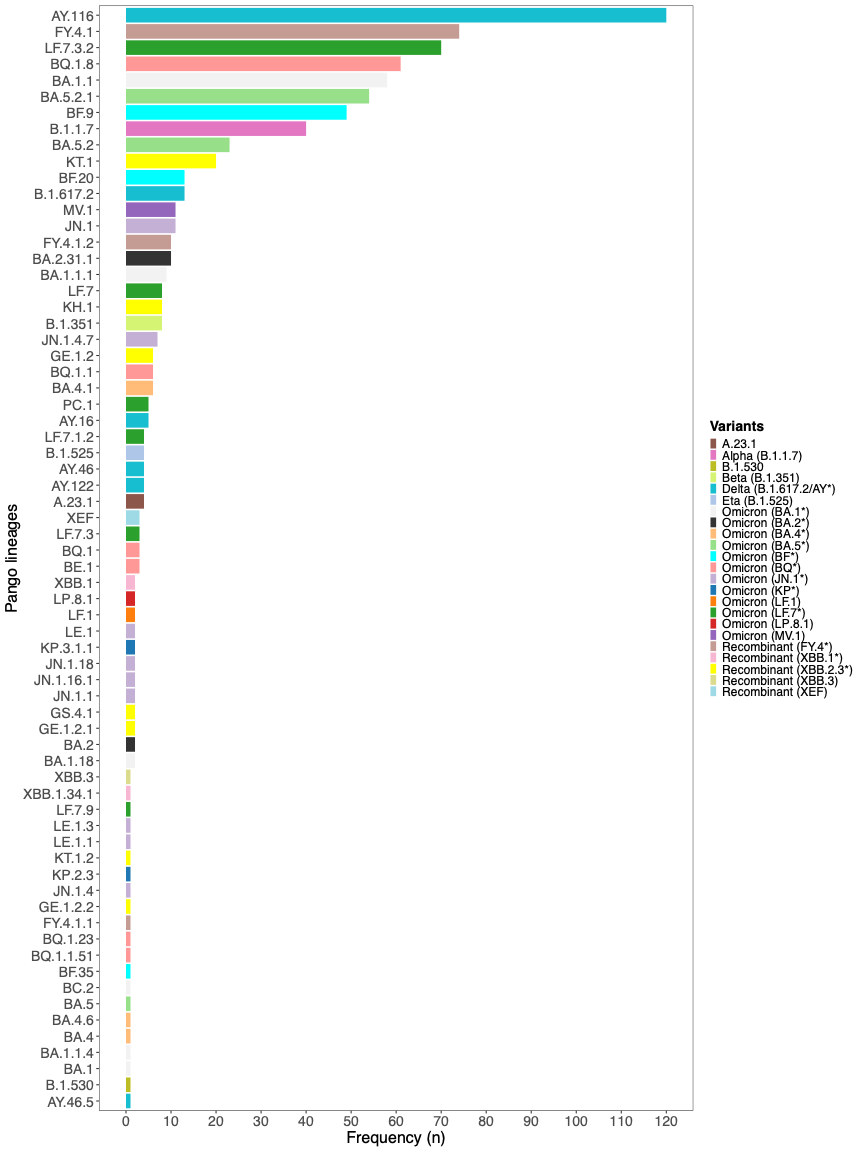

#### Supplementary Figure 1: Frequency of detected SARS-CoV-2 lineages detected between December 2020 and February 2025 in Kilifi Kenya from SARS-CoV-2 cases presenting with ARI symptoms in five outpatient healthcare facilities.

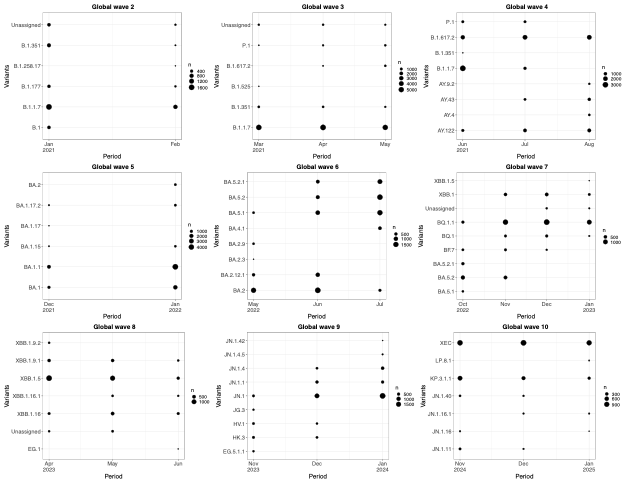

#### Supplementary Figure 2: Temporal patterns of monthly top five PANGO lineages globally from sequences deposited in GISAID across different time intervals of waves observed in Kilifi Kenya between December 2020 and February 2025.

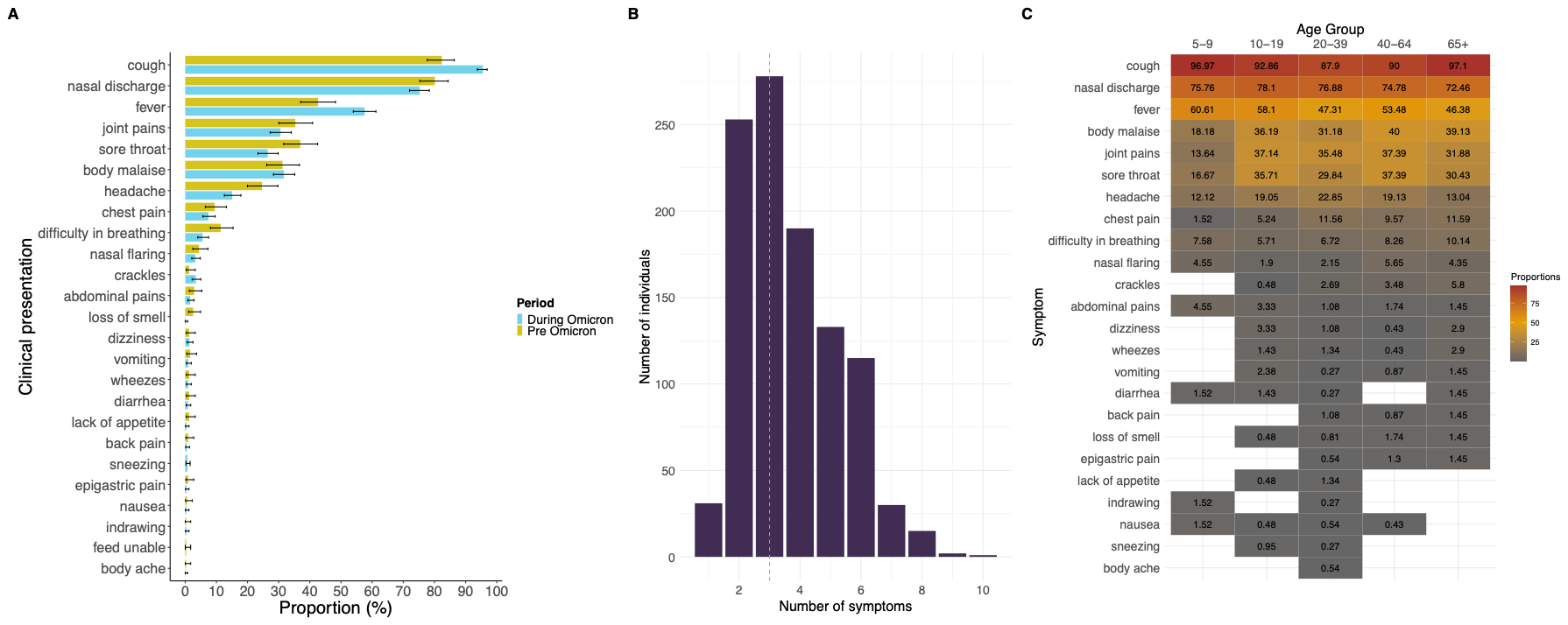

#### Supplementary Figure 3: Symptom epidemiology in SARS-CoV-2 positive individuals seeking outpatient care. **(A)** Prevalence of COVID-19 symptoms in the pre-Omicron and Omicron periods. Error bars indicate 95% credible intervals (Cis). **(B**) Frequency of symptoms reported per individual. The dotted line shows the median number of symptoms reported per individual. **(C)** Prevalence of COVID-19 symptoms by age. A cut-off of five years and above was used to avoid bias among the symptoms as some cannot be recorded for children (e.g., headache). A gradient colour scale is used to show high and low proportions.
